# Airway-specific autoantibodies identify a subset of patients with fibrotic interstitial lung disease

**DOI:** 10.1101/2021.07.29.21261213

**Authors:** Karim Boustani, Poonam Ghai, Rachele Invernizzi, Richard J. Hewitt, Toby M. Maher, Quan-Zhen Li, Philip L. Molyneaux, James A. Harker

## Abstract

**Background:** Fibrotic interstitial lung disease (fILD) has previously been associated with the presence of autoantibody. While studies have focused on systemic autoimmunity, the role of local autoantibodies in the airway remains unknown. We therefore extensively characterised the airway and peripheral autoantibody profiles in patients with fILD and assessed association with disease severity and outcome.

**Methods:** Bronchoalveolar lavage (BAL) was collected from a cohort of fILD patients and total airway antibody concentrations were quantified. An autoantigen microarray was used to measure IgG and IgA autoantibodies against 124 autoantigens in BAL from 40 idiopathic pulmonary fibrosis (IPF), 20 chronic hypersensitivity pneumonitis (CHP), 20 connective tissue disease-associated ILD (CTD-ILD) patients and 20 controls.

**Results:** A subset of patients with fILD but not healthy controls had a local autoimmune signature in their airways that was not present systemically, regardless of disease. The proportion of patients with IPF with a local autoantibody signature was comparable to that of CTD-ILD, which has a known autoimmune pathology, identifying a potentially novel subset of patients. The presence of an airway autoimmune signature was not associated with reduced survival probability or changes in lung function in the cohort as a whole. Patients with IPF had increased airway total IgA and IgG1 while subjects with CHP had increased airway IgA, IgG1 and IgG4. In patients with CHP, increased airway total IgA was associated with reduced survival probability.

**Conclusion:** The presence of airway autoantibodies identifies a unique subset of patients with fILD and the mechanisms by which these autoantibodies contribute to disease requires further investigation.

## Introduction

Interstitial lung disease (ILD) is an umbrella term for a group of devastating, chronic lung diseases including idiopathic pulmonary fibrosis (IPF), chronic hypersensitivity pneumonitis (CHP) and connective tissue disease-associated ILD (CTD-ILD)^1^. Each of these diseases has a unique aetiology and, in the case of IPF which is the most common form of ILD, the cause remains unknown. This is a growing problem in the UK, with more than 5,000 cases diagnosed annually and despite therapy a 5-year survival of approximately 20%^2^. Despite differing aetiologies and triggers there is growing evidence of shared pathogenesis across the spectrum of fibrotic interstitial lung disease (fILD). These diseases arise in response to microinjuries to the respiratory epithelium, which trigger an aberrant wound healing response in genetically susceptible aged individuals. The immune system is known to play a role in the pathogenesis of both CHP, largely driven by known environmental antigens, and CTD-ILD, driven by self-antigens. The role of the immune system in the pathogenesis of IPF is however less clear. Recent work has highlighted changes in the lung immune response, especially within the macrophage population, and their correlation with disease outcome, but a source of antigen stimulation has yet to be identified^3^.

Autoimmunity occurs as a result of a breakdown of tolerance within the adaptive immune cell compartment, resulting in the generation of antibodies by plasma cells which target self-antigens. The presence of circulating autoantibodies is used to diagnose CTD-ILD clinically. These circulating autoantibodies are often against nuclear components such as Ro, La, Jo1, Scl70 and double-stranded DNA (dsDNA)^4–6^. Associations between circulating autoantibodies and disease outcome are not however limited to CTD-ILD, and other forms of fILD, such as CHP as well as other chronic lung diseases such as chronic obstructive pulmonary disease (COPD) have been shown to have similar associations^6–10^. Supporting a role for antibody mediated immunity in the pathogenesis of some patients with pulmonary fibrosis, Rituximab, a monoclonal antibody that specifically depletes B cells, has also been shown to be beneficial in a subset of patients with either CTD-ILD or CHP^11–13^. Much less is known, however, about the contribution of the antibody response to IPF, although a small scale trial of rituximab in combination with plasmapheresis showed some benefit during acute exacerbations of IPF^14^. Further evidence for humoral dysregulation being a contributor to disease pathology comes from the identification of multiple circulating autoantibodies against various components of alveolar epithelial cells, extracellular matrix components such as periplakin and vimentin, and other lung-specific proteins such as BPIFB1^5–7, 15–20^. It is not well understood how these autoantibodies drive pathology but autoantibodies targeting proteins expressed almost exclusively in the lung parenchyma such as KCNRG and BPIFB1 have been identified in patients with fibrotic lung disease^18, 21, 22^. This suggests that autoantibodies against proteins expressed in lung tissue may induce damage and, subsequently, inflammation and aberrant wound repair resulting in fibrosis.

In this study, we sought to determine whether there is a local autoimmune signature in the airways of patients with fILD and whether the presence of airway autoantibodies could be used to predict disease outcome. We demonstrate that there is a significant increase in airway IgA, IgG1 and IgG4 in IPF, and increased IgA and IgG4 in CHP. A subset of patients with fILD had distinct airway autoantibody profiles, the presence of which is not associated with autoantibodies in the circulation. These findings present a novel observation that there is a distinct airway autoantibody signature in a subset of patients with fILD that is not present in healthy individuals. Further work will be required to assess the implications of these findings in clinical practice and elucidate the pathological role airway autoantibodies may play.

## Methods

### Patients and sampling

Patients undergoing routine diagnostic bronchoscopy at the Interstitial Lung Disease Unit in the Royal Brompton Hospital between May 2014 and December 2019 were recruited prospectively. The study was approved by the East London and the City Research Ethics Committee (15-LO-1399) and the South Central Hampshire Research Ethics Committee (15/SC/0101). Written informed consent was obtained from all subjects. Subjects with histories of upper or lower respiratory tract infections, antibiotic use in the prior three months or history of acute exacerbations were excluded. Bronchoscopy with bronchoalveolar lavage (BAL) was performed as described previously^23, 24^.

### BAL processing

BAL samples were passed through a 70µm sterile strainer and centrifuged (700*g*; 5 min; 4°C). Supernatants were stored at −80°C for further use.

### ELISAs

Total antibody ELISAs for IgA, IgG, IgM, IgG1, IgG2, IgG3 and IgG4 were carried out on BAL supernatants according to manufacturer’s instructions (Invitrogen, UK). Vitronectin and Collagen V-specific ELISAs were developed based on a standard sandwich ELISA assay. Plates were coated with either 0.625µg/ml of recombinant human vitronectin (PromoCell, Germany) or 0.1µg/ml of human collagen V (Sigma-Aldrich, UK). Serial dilutions of BAL and plasma were performed starting at neat for BAL and 1:20 for plasma. Plates were incubated with HRP-conjugated anti-human IgG or IgA antibodies at a 1:4000 dilution (SouthernBiotech, USA) and developed with TMB. Absorbance at 450nm was measured using a SpectraMax i3x plate reader (Molecular Devices, USA).

### Autoantigen array

IgA and IgG autoantibody reactivities against a panel of 124 autoantigens were measured in 50µl of BAL using an array developed by the University of Texas Southwestern Medical Center^8, 25^. Briefly, samples were incubated with autoantigens on microarray slides before being incubated with secondary fluorophore-conjugated anti-human IgG or IgA antibodies. Following detection, images were analysed by Genepix Pro 6.0 (Molecular Devices, USA). Average net fluorescence intensities (NFI) and signal-to-noise ratios (SNR) were calculated by subtracting negative control (PBS) fluorescence values from sample fluorescence values.

### Statistical analysis

Statistical analyses were performed using Prism software (GraphPad Software). Kaplan-Meier analysis was used to compare mortality between subjects with or without airway autoantibodies. Both univariate and multivariate Cox Proportional Hazard regression analyses were carried out in R. Heatmaps were generated using Morpheus (Broad Institute, MA, USA).

## Results

### IgA, IgG1 and IgG4 antibodies are increased in IPF and CHP airways

Given that previous studies have shown alterations in both circulating and airway total antibody in fILD, we first investigated whether antibody concentrations were elevated in patients with IPF and CHP compared to healthy controls^15^. A significant increase in serum IgG but not IgA or IgM was observed in patients with IPF compared to healthy controls (Figure 1Ai). In the BAL we observed a statistically significant increase in IgA in both patients with IPF and CHP compared to healthy controls, as well as an increase in IgG in patients with CHP (Figure 1B). There also appeared to be a trend toward increase in IgG in patients with IPF compared to healthy controls (Figure 1B). Analysis of IgG isotypes showed that IgG1 and IgG4 concentrations were significantly increased in patients with CHP and IgG1 was also significantly increased in patients with IPF compared to healthy controls (Figure 1C). Taken together, these data suggest that increases in airway IgG and IgA are features of both IPF and CHP.

**Figure 1.**
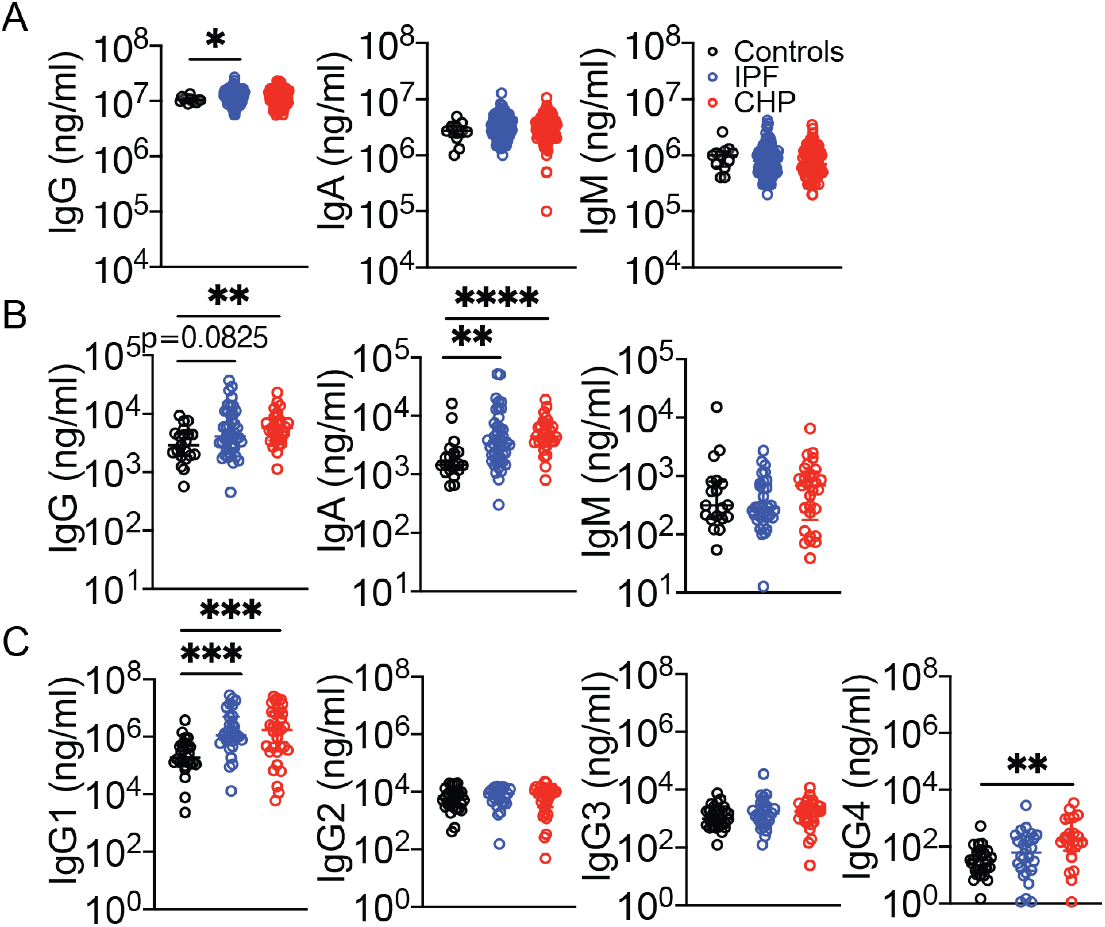
Local antibodies are increased in patients with IPF and CHP. **(A)** Total IgG, IgA and IgM concentrations in plasma from patients with IPF and CHP, and healthy controls. **(B)** Total IgG, IgA and IgM concentrations in BAL from patients with IPF and CHP, and healthy controls. **(C)** IgG1, IgG2, IgG3 and IgG4 concentrations in BAL from patients with IPF and CHP, and healthy controls. Each point represents an individual subject. Error bars represent median and interquartile range. Data were analysed using a non-parametric Kruskal-Wallis test with Dunn’s multiple comparison. *, 0.05; **, p<0.01; ***, p<0.001; ****, p<0.0001.

We performed additional analysis to determine whether increases in airway antibodies are associated with disease severity and predicted survival. Although there were weak negative correlations between airway antibody and lung function in patients with IPF and CHP, this did not influence predicted survival (Figures S1A, S1B, S1C). However, in patients with CHP, higher airway IgA concentrations were associated with significantly reduced survival probability (*p*=0.0082) (Figure S1D). Collectively, these data suggest that airway antibody concentrations may be associated with poorer disease outcome in some forms of fILD.

### A distinct autoimmune signature is present in a subset of patients with fILD

Because IgA and IgG were increased in the airways, we hypothesised this could be linked to increased IgA and IgG autoantibodies in the airways of patients with fILD. Hence, we performed IgG and IgA autoantibody arrays on 100 BAL samples, which included patients with IPF (n=40), CHP (n=20) and CTD (n=20) as well as healthy and non-disease controls (n=20). Demographic and clinical features are shown in Table 1. Notably, we were able to identify two distinct subsets of patients, those with detectable airway IgG (Figure 2A) and IgA (Figure 2B) autoantibodies and those without by hierarchical clustering. Within fILD patients who were positive for airway autoantibodies, we were able to identify IgG and IgA autoantibodies against a range of nuclear proteins (e.g. dsDNA, chromatin, nucleosome, POLB, ssRNA and ssDNA), extracellular matrix-associated (e.g. Collagen I-VI, heparin and elastin) and epithelial-associated proteins (e.g. vimentin and vitronectin) (Figures 2A; 2B). IgG and IgA autoantibodies were also detectable in two non-disease controls, however one of these was a patient with COPD and the other was a patient with bronchiectasis (Figures 2A; 2B). Both of these conditions have been previously associated with presence of airway autoantibodies and those samples taken from healthy patients had no detectable autoantibodies^8, 9, 26–28^.

**Table 1.** Patient demographics for autoimmune array.

| **Parameter** | **Controls (n=20)** | **IPF (n=40)** | **CHP (n=20)** | **CTD (n=20)** |
| --- | --- | --- | --- | --- |
| Age, year mean (±SD) | 64 (±9) | 73 (±6)<sup>***</sup> | 71 (±8) | 70 (±12)<sup>*</sup> |
| Female sex, n (%) | 25 | 33 | 60 | 50 |
| Ex-smoker or current smoker, n (%) | 56 | 68 | 40 | 53 |
| FEV1, % predicted (±SD) | 94 (±19) | 92 (±21) | 83 (±21) | 86 (±28) |
| FVC, % predicted (±SD) | 101 (±15) | 91 (±21) | 87 (±22) | 82 (±24)<sup>*</sup> |
| DLCO, % predicted (±SD) | 81 (±29) | 55 (±15) | 60 (±16) | 50 (±19)<sup>*</sup> |

FEV1: Forced expiratory volume in one second
FVC: Forced vital capacity
DLCO: Diffusing capacity of the lungs for carbon monoxide
Mann-Whitney U test: \*, $p<0.05$ ; \*\*, $p<0.01$ ; \*\*\*, $p<0.001$ ; \*\*\*\*, $p<0.0001$ .

**Figure 2.**
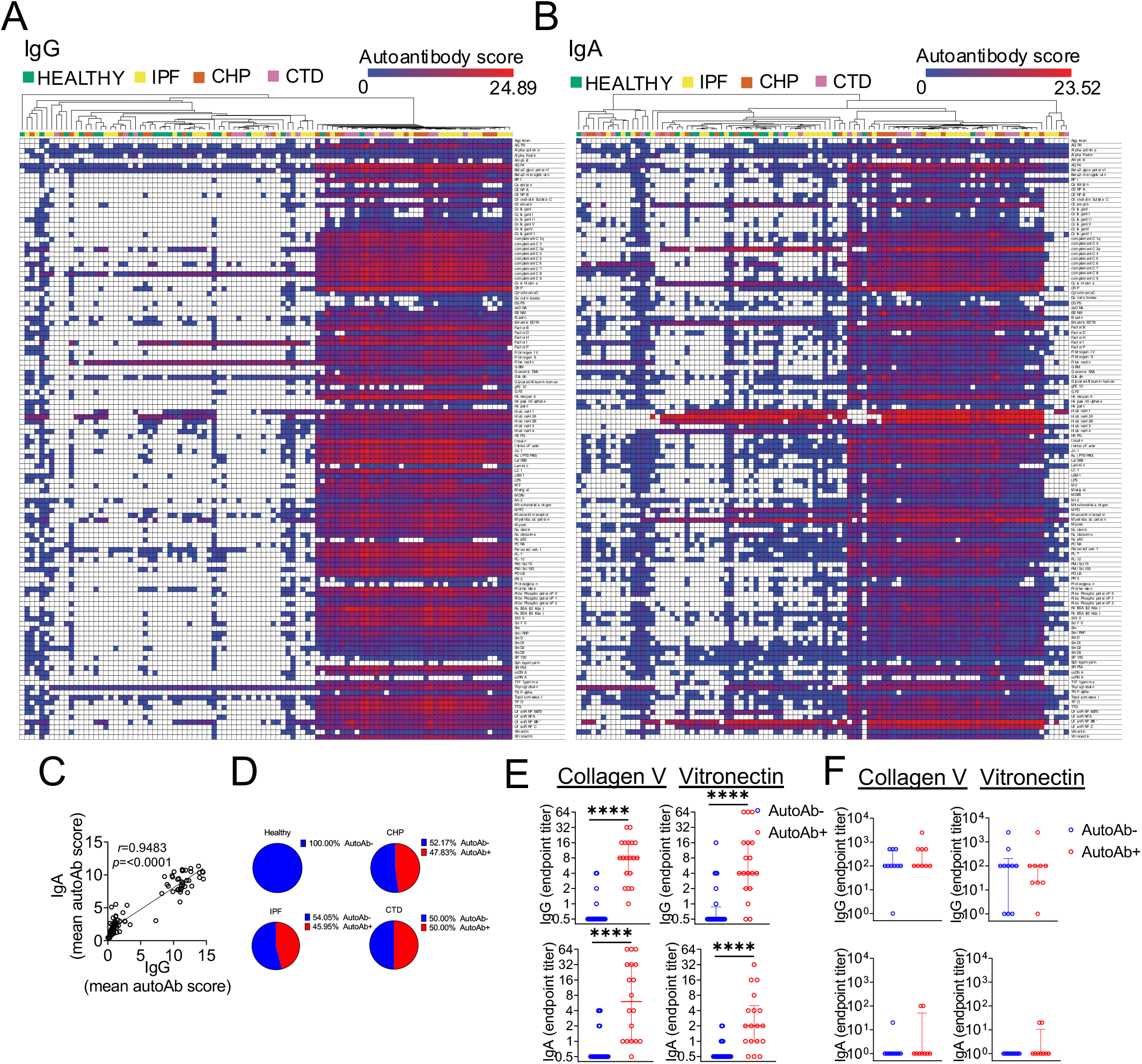
IgG and IgA autoantibodies are detectable in the airways of a subset of patients with fILD. BAL IgG **(A)** and IgA **(B)** autoantibodies were screened for reactivity against 98 self antigens in IPF (n=40), CHP (n=20) or CTD patients (n=20) and non-fibrotic controls (n=20). Heatmaps show autoantibody scores, calculated as log2 transformed(net fluorescence intensity x signal-to-noise ratio+1). White squares indicate samples for which autoantibody signal was not detectable against specific autoantigens. Samples were clustered hierarchically. **(C)** Pearson correlation between mean IgG and IgA autoantibody scores. **(D)** Proportions of autoantibody positive individuals in each study group. **(E)** BAL from the array was used to validate the findings independently in a sandwich-ELISA assay in order to validate presence of autoantibodies against collagen V and vitronectin. **(F)** Serum dilutions were also analysed for autoantibodies against collagen V and vitronectin. Each point in (C), (E) and (F) represents an individual subject. Error bars represent median and interquartile range. Data in (E) and (F) were analysed using a non-parametric Mann-Whitney U test. *, p<0.05; **, p<0.01; ***, p<0.001; ****, p<0.0001.

We next asked whether the patients with increased IgG autoantibodies were also those with increased IgA autoantibodies. We observed a statistically significant correlation between IgG and IgA autoantibody scores (Figure 2C). Approximately half of the patients with IPF, CHP and CTD had a detectable autoantibody signature in their airways (Figure 2D).

Patients who tested positive in the autoantigen array for antibodies against collagen V and vitronectin, both of which are expressed in lung tissue, also had significantly increased titers of anti-collagen V and anti-vitronectin autoantibodies in the airways when determined independently by ELISA (Figures 2E; S2A)^29, 30^. Importantly, no difference was observed in serum titers of anti-collagen V and anti-vitronectin antibodies between the AutoAb- and AutoAb+ patients or healthy controls, suggesting that the presence of autoantibodies in these patients was unique to the airways (Figures 2F; S2B).

### Presence of lower airway autoantibodies is not associated with disease progression and survival

Finally, we wanted to determine whether the presence of airway autoantibodies is associated with more severe disease. Principal components analysis with k means clustering based on IgG autoantibody scores was performed on all patients with fILD but not healthy controls and identified 3 distinct clusters of patients (Figure 3A). Clusters 1 and 2 corresponded to subjects who had positive autoantibody signal (Figure 3A). There was no difference in lung function between any of the clusters (Figure 3B). Cluster 3 contained a higher percentage of progressors, as defined by patients who had a decline in FEV1 greater than 10% (Figure 3C). Cluster 1 had an increased percentage of patients with IPF (56.25%) and CTD (25%) compared to clusters 2 and 3 while clusters 2 and 3 had increased proportions of patients with CHP (40.1% and 33.3% respectively) (Figure 3D). Overall, there was no difference in survival probability between any of the clusters (Figure 3E).

**Figure 3.**
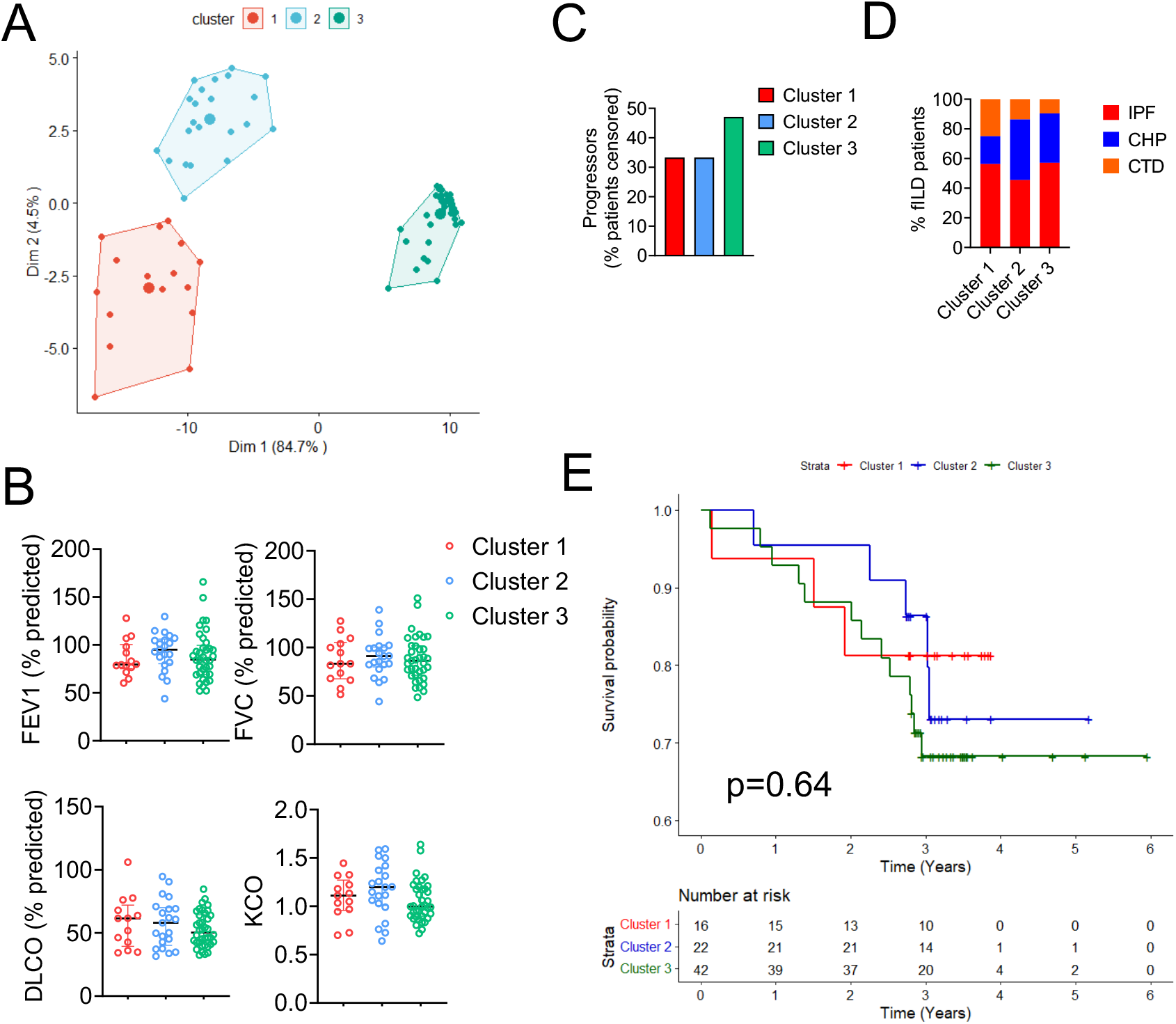
Presence of airway autoantibody in patients with fILD is not associated with reduced lung function or survival probability. (A) Principal components analysis followed by k means clustering based on airway IgG autoantibody scores. (B) Patient lung function scores by cluster. (C) Percentage of patients who show lung function decline as a proportion of all patients censored by cluster. (E) Kaplan-Meier curve generated using the Cox proportional hazards model showing predicted survival probability in patients in each of the clusters. Each point in (A) and (B) represents an individual subject. Error bars represent median and interquartile range.

To identify the top putative autoantigens responsible for variance in PC1, percentage contributions were analysed. Fibrinogen IV, Fibrinogen S, Ribophosphoprotein P1, PM.Scl.75, Vitronectin and RoSSA accounted for the highest contribution to variance in PC1 (Figure S3A). Stratification of patients based on presence or absence of autoantibodies against these antigens showed no difference in survival probability (Figure S3B). Similarly, for PC2, chondroitin sulfate C, nup 62, heparin, ssRNA, decorin bovine and collagen II had the highest contribution to variance (Figure S3C). Stratification of patients based on presence or absence of these autoantibodies show no significant difference in survival probability (Figure S3D). Taken together, these data suggest that while the airways of approximately half of fILD patients are enriched for autoantibodies against cellular and nuclear antigens, these do not associate with disease severity and outcome.

## Discussion

The presence of circulating autoantibodies as well as their impact on disease outcome in patients with interstitial lung disease has been widely reported. Here we sought to address whether patients with interstitial lung disease have a local airway autoimmune signature and whether this could be used as a predictor of disease severity and outcome. We show that IgA, IgG1 and IgG4 are increased in the airways of patients with fILD and identify a subset of patients with local but not systemic autoantibodies.

Although patients with IPF had modest increases in serum IgG, patients with IPF and CHP both had increases in airway IgG and IgA. In agreement with a previous study, total airway antibody was not associated with worse predicted survival^15^. Specifically, IgG1 was increased in IPF airways while IgG4 was increased in IPF and CHP airways. Although IgG is predominantly involved in systemic responses, it is also the most abundant antibody class in the lower respiratory tract^31, 32^. Unlike IgA and IgM, both of which are transcytosed via pIgR, IgG is transcytosed bidirectionally via the action of the neonatal Fc receptor (FcRn)^33^. IgG1 is involved in pro-inflammatory responses including complement activation while IgG4 suppresses immune activation and promotes tolerance^34^. IgA was also increased in IPF and CHP airways. In the gut, IgA is responsible for maintaining homeostatic relationships with commensals in the GI tract through immune exclusion^35^. However, its role in the lower respiratory tract remains to be defined. In the gut, both IgG1 and IgA play important roles in maintaining commensals^31, 34^. It has previously been reported that patients with IPF and CHP have increases in lower respiratory tract bacterial burdens^23, 36–38^. It is therefore possible that the increase in local IgA and IgG1 in the airways of fILD patients are a compensation mechanism for the enhanced bacterial burden observed in these patients.

Over the last decade, studies have begun to explore the role of autoantigens in fILD pathology. However, the relationship between systemic autoimmune disease and interstitial lung disease remains poorly understood. One of the first circulating epithelial autoantibodies to be identified in pulmonary fibrosis was anti-cytokeratin 8 and this provided evidence to support the hypothesis of autoantibody-mediated lung injury^39^. Since then, multiple circulating autoantibodies have been reported against human epithelial and extracellular matrix proteins including annexin 1, heat shock protein 70, vimentin and periplakin, some of these correlating with disease severity^4, 5, 15, 20, 40^.

We confirm and extend previous analyses of autoantibodies in fILD and show for the first time by large-scale array that autoantibodies against a range of matrix components are also detectable locally within the airways of a subset of patients with fibrotic lung disease. This data suggests that there is a specific local autoimmune signature in some patients with fibrotic lung disease. Importantly, these data also support a link between autoimmunity and tissue damage and fibrosis. Early studies showed that patients with fibrosing alveolitis have increased concentrations of IgG autoantibodies to proteins expressed within lung tissue^41^. Shum and colleagues elegantly showed that autoantibody targeting of BPIFB1, a protein expressed exclusively in the lung tissue, results in the development of lung fibrosis in a murine model^18, 22^.

Here, we show that patients have autoantibodies against a range of proteins expressed within the lung and some which may be associated with tissue damage, such as nuclear proteins. Indeed, autoantibodies against some of the antigens in our array such as vimentin have been previously reported in fILDs^4, 16^. This broad pattern of autoantibody specificity has also been reported in the serum from severe COPD patients and was particularly associated with emphysema, suggesting that autoantibody profiles differ between disease groups^8^. Interestingly, most of the autoantibodies we present here are against proteins that are often upregulated in fILD. Vitronectin is significantly increased in the BAL of patients with ILD^30^. Similarly, many of the components of the extracellular matrix such as the collagen proteins, particularly collagen I and V are overexpressed in the wound repair process. Collagen V plays a central role in the formation of fibrillar collagen mesh and is also involved in regulating the fibre size^42^. In parenchymal fibrosis, collagen V is highly overexpressed in IPF lungs and much of the morphological disorganisation of fibrillar collagen can be attributed to this aberrant expression of collagen V^19, 42^. Interestingly, it has been reported that 40-60% of patients with IPF have autoimmune responses against collagen V, similar to the proportions observed in the current study^19^. It is possible that increased expression of collagen V in lung fibrosis results leads to an aberrant humoral response against an otherwise innocuous protein. A recent phase I clinical trial involving oral immunotherapy with bovine collagen V in patients with IPF led to stabilised of lung function and reduced matrix metalloproteinase expression, possibly through the induction of humoral tolerance^29^.

The site of autoantibody production in fILD remains elusive. The presence of a local airway but not systemic autoimmune signature suggests that the production of autoantibodies may be occurring locally. Early studies by Rangel-Moreno and colleagues showed that autoantibody production can occur locally within lymphocyte aggregates within fibrotic lung tissue^43^. Specifically, they showed that autoantibodies against both vimentin and citrullinated proteins localise around B cell aggregates within the fibrotic lung tissue^16, 43^. Indeed, B cell aggregates are observed in biopsy tissue from human IPF lungs^44^. Local autoantibody production is also supported by tissue-resident B cells in other organs such as the upper respiratory tract and synovia in chronic rhinosinusitis with nasal polyps and rheumatoid arthritis, respectively^45, 46^. It is important to note that lymphoid aggregates are also not observed in all patients with RA-associated lung fibrosis^43^. This may explain why only some patients develop a local autoimmune signature. However, our findings suggest that the link between local autoantibody and disease progression cannot be assumed and may also depend on other factors such as time of sampling. One of the main limitations of the study is that the median follow-up time is 3 years and that may limit our ability to detect differences in survival between those patients with and without local autoantibodies. However, there is no signal of rapid deterioration.

We have demonstrated for the first time that there is a broad spectrum of autoantibodies present locally within the airways of patients with fILD that is not present in the circulation, supporting our hypothesis that local antibody-mediated tissue damage can drive pathology of lung fibrosis. Subsequent investigations should therefore aim to define the role of these autoantibodies in tissue damage. Ultimately, if humoral dysregulation is a key player in severe disease then a number of targeted therapies may be more beneficial to outcome than non-specific treatments.

## Data Availability

N/A

## Acknowledgements

This work was funded by a Rosetrees Seed Fund to PLM and JAH (A2172). KB was funded by an Asthma UK studentship to JAH and PLM as part of the Asthma UK centre in Allergic Mechanisms of Asthma (AUK-BC-2015-01).

---

**Figure S1.**
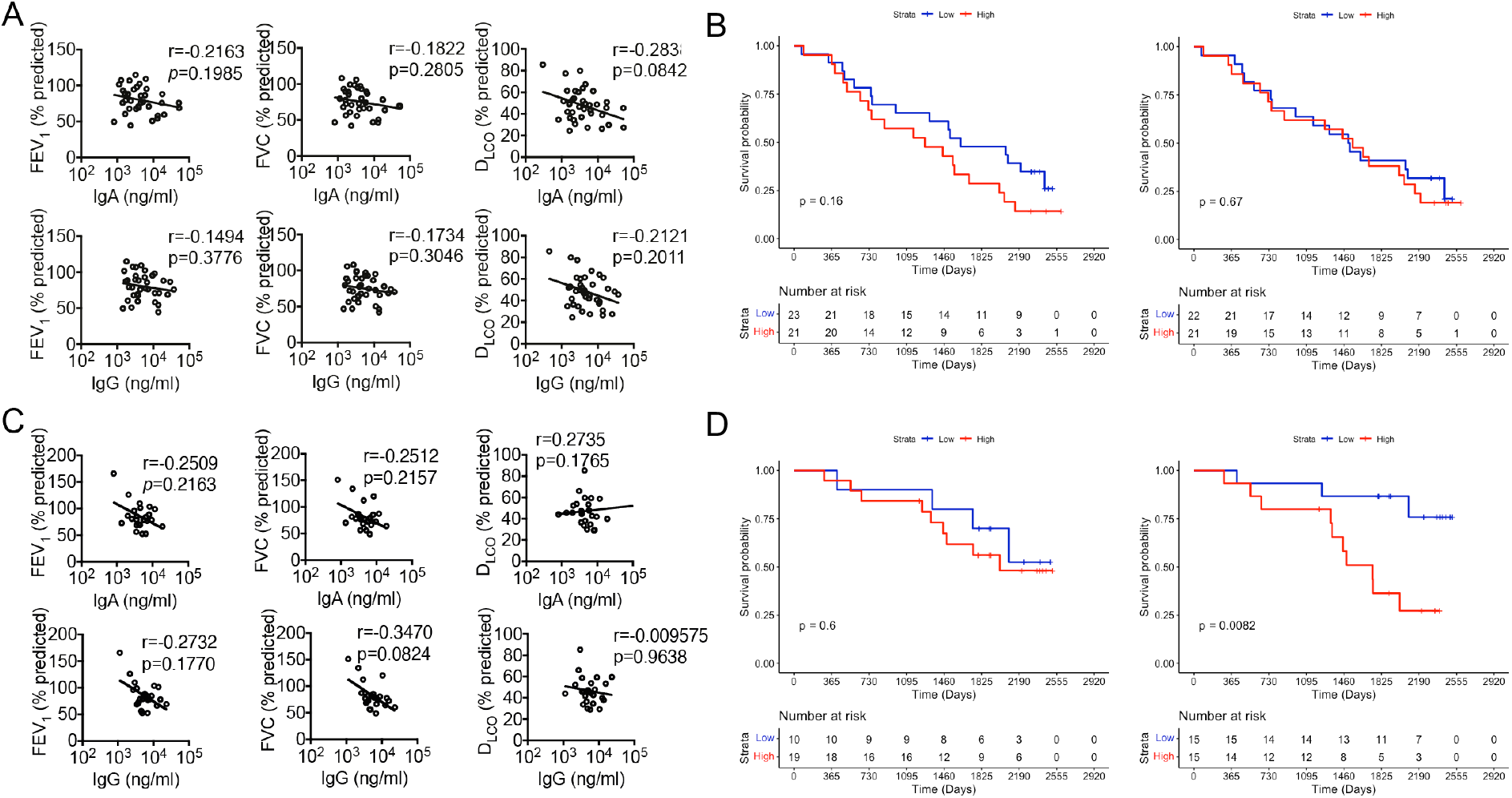
High airway IgA concentrations are associated with reduced survival probability in patients with CHP. **(A)** Pearson correlations between lung function measurements and airway antibody concentrations in BAL from patients with IPF. **(B)** Kaplan-Meier curve generated using the Cox proportional hazards model showing predicted survival probability in patients with IPF dichotomised by median concentrations of IgG and IgA. **(C)** Pearson correlations between lung function measurements and airway antibody concentrations in BAL from patients with CHP. **(D)** Kaplan-Meier curve generated using the Cox proportional hazards model showing predicted survival probability in patients with CHP dichotomised by median concentrations of IgG and IgA. FEV1: forced expiratory volume in 1 second; FVC: forced vital capacity; DLCO: diffusing capacity for carbon monoxide.

**Figure S2.**
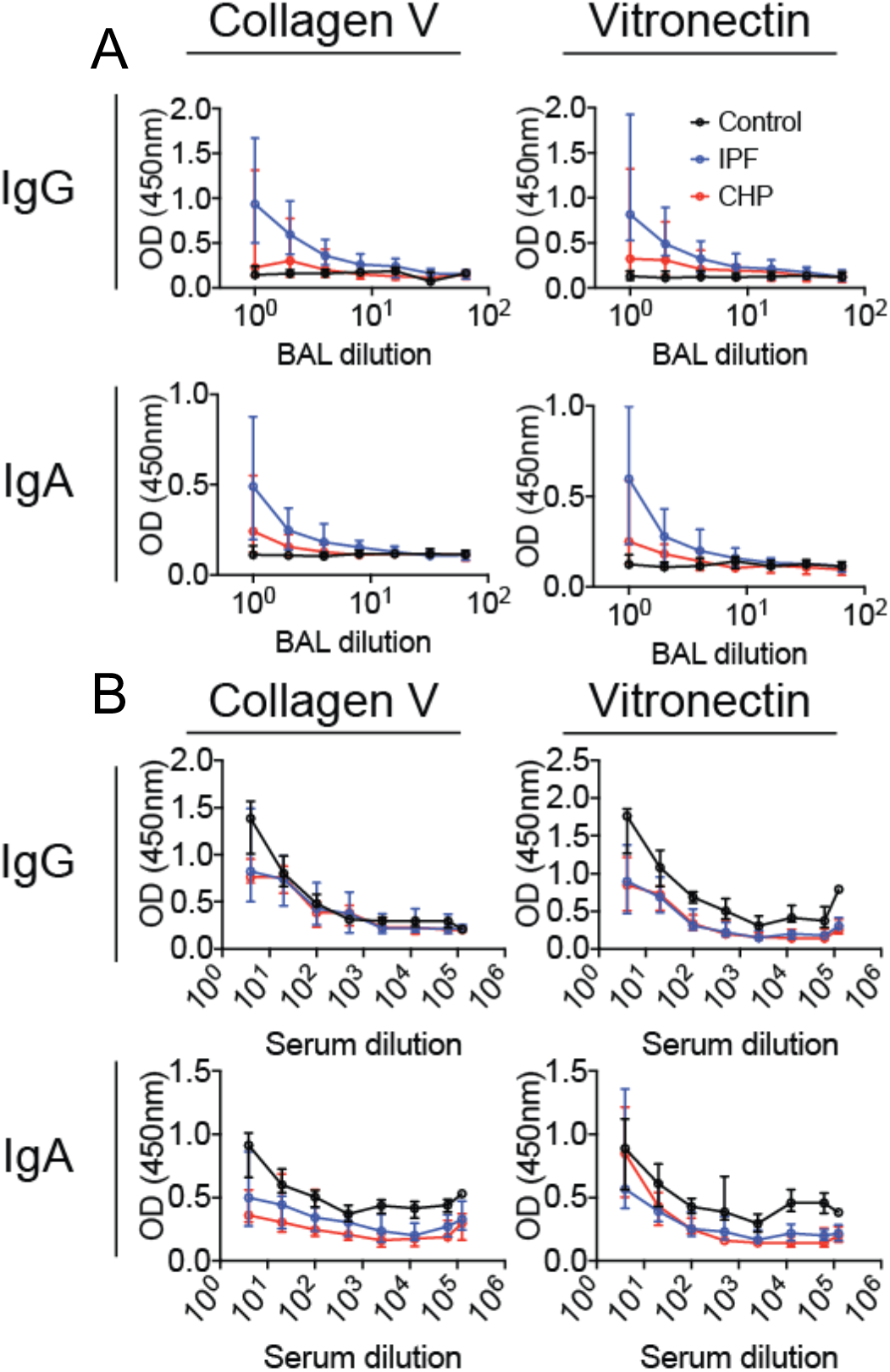
Dilution curves for BAL and serum. Serial dilutions of BAL **(A)** and serum **(B)** were analysed for autoantibodies against human collagen V and vitronectin. Each point represents the median optical density (OD) for each disease group. In BAL: control (n=13), IPF (n=15), CHP (n=14); in serum: control (n=6), IPF (n=15), CHP (n=10). Error bars represent median and interquartile range. Data were analysed using a two-way ANOVA.

**Figure S3.**
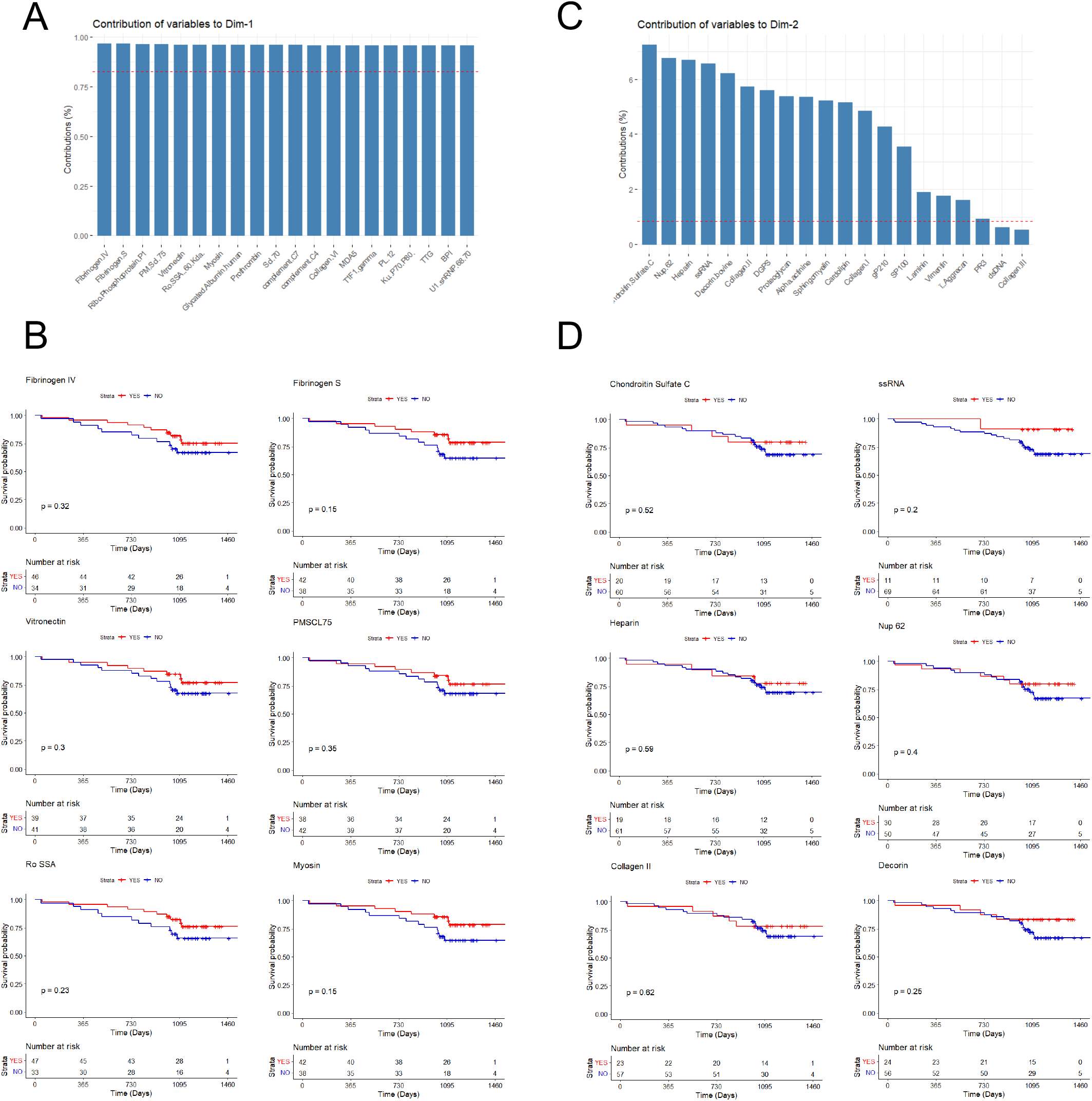
Specific autoantibodies driving patient clustering do not associate with survival. (A) Top 20 autoantigens contributing to PC1. (B) Kaplan-Meier analysis of top 6 proteins driving variance in PC1. (C) Top 20 autoantigens contributing to PC2. (D) Kaplan-Meier analysis of top 6 proteins driving variance in PC1. Kaplan-Meier curve generated using the Cox proportional hazards model showing predicted survival probability in patients in each of the clusters.

